# Analysis of cardiomyocyte nuclei in human cardiomyopathy reveals orientation dependent defects in shape

**DOI:** 10.1101/2020.08.14.20168310

**Authors:** Daniel Brayson, Elisabeth Ehler, Cristobal G. dos Remedios, Catherine M. Shanahan

## Abstract

Cardiomyopathies are progressive diseases of heart muscle often caused by mutations in genes encoding sarcomeric, cytoskeletal and nucleoskeletal proteins though in many cases the cause of disease is not identified. Whilst nucleus hypertrophy has been described, it is not known whether nucleus shape changes are a general feature of cardiomyopathy. Due to the rod-shaped nature of cardiomyocytes and their elliptical nuclei we hypothesised that orientation of analysis would be an important determinant of any changes observed between patients exhibiting primarily unexplained cardiomyopathy and control samples from non-failing donors. To investigate this we performed image analysis of cardiomyocyte nuclei in myocardial cryosections from a cohort of cardiomyopathy patients. We discovered that circularity, solidity and aspect ratio were sensitive to orientation of the myocardium and that in the transverse plane only circularity was reduced in cardiomyocyte nuclei of cardiomyopathy patients. These findings show that orientation dependent changes in nucleus shape may be a property of cardiomyopathy and with appropriate follow up studies, may prove to have mechanistic and diagnostic value.

## Introduction

Cardiomyopathies are diseases of cardiac muscle causing cell and tissue level deficiencies that precede a progressive decline in the ability of the heart to contract and relax, resulting in heart failure and sudden cardiac death (1). In many cases a root cause of disease is identifiable by performing genetic testing for known disease-causing gene variants and by studying patient and family history (2). However, unexplained cardiomyopathy in which a specific cause is not discernible may account for up to 50% of all cardiomyopathies (3), meaning there is a need to better understand disease processes of these so called ‘idiopathic’ cardiomyopathies (4). Investigating the cardiomyocyte nucleus may present an opportunity to achieve some understanding. For example, the number of sets of chromosomes, or ploidy, in cardiomyocyte nuclei has been shown to increase with age and may relate to a reduced intrinsic regenerative capacity (5). Accordingly, this may be linked to cardiomyocyte nucleus size since nucleus hypertrophy has been observed in explanted tissue from dilated (DCM) and hypertrophic (HCM) cardiomyopathies, reflecting ‘increased biosynthetic processes’ such as DNA replication and DNA damage repair (6). These lines of evidence point towards important roles for nucleus size in disease. In contrast, nuclear morphology defects, where the shape of the nucleus becomes distorted, have not been investigated in the broad context of cardiac muscle disease, whereas they are well established as a cardinal feature of laminopathic diseases such as progeria (7, 8). The gene responsible for the majority of laminopathies is the *LMNA* gene encoding the nuclear filament proteins lamins A and C, which arise as a result of alternate splicing. Lamins A and C comprise the bulk of the nucleoskeleton, facilitating a mechanical connection to the cytoskeleton along with Sad1 and UNC (SUN) proteins and nuclear envelope spectrin repeat proteins (nesprins) as part of the linker of nucleo-cytoskeleton (LINC) complex (9). Importantly, the *LMNA* gene is also a candidate gene for DCM potentially accounting for 6-8 % of familial DCM cases (10, 11). We recently discovered a specific role for the accumulation of the lamin A precursor, prelamin A, in DCM and found disruption to nuclear morphology, inflammation and senescence (12). Cardiomyocyte nuclear morphology changes have also been described in other *in vitro* and *in vivo* systems investigating LINC complex dysfunction (13, 14) and cytoskeletal disruption (15-17). These studies mostly performed analysis of nuclear morphology on cardiomyocytes *in vitro*, isolated from the organ and laid out flat in the longitudinal axis. Although isolation of human cardiomyocytes is technically possible (5), due to the complexities associated with heart transplant such as logistics and timing, diseased tissues explanted from patients are often cryopreserved or paraffin fixed and embedded in epoxy resin, and heart sections are often the most readily available sample type available to investigators. Therefore, analysing nucleus morphology in cryopreserved or paraffin embedded heart sections may represent an additonal opportunity to gain insight into nuclear morphology of cardiomyopathy versus non-failing (NF) left ventricle control samples from donor heart tissue, and examine whether it is affected in hearts in the latter stages of cardiomyopathic heart failure. Since cardiomyocyte nuclei are elliptical, we hypothesised that axis orientation/plane of view would be an important determinant of any outcome derived from performing such analyses.

## Results & Discussion

To investigate the importance of nucleus orientation, paired image analysis of size parameters in heart cryosections from DCM and NF control samples displaying cardiomyocytes in the longitudinal axis (Fig. 1A) versus the transverse plane (Fig. 1B) showed that groupwise; area, perimeter, length and Feret’s diameter (defined as the distance between the two parallel planes restricting an object perpendicular to that direction) were significantly lower in the transverse axis for both sample sets (Figs. 1C-F). Nuclear width was similar between both analyses in both groups (Fig. 1G). Overall, shape characteristics (defined in detail in the methods) displayed more nuanced changes. Aspect ratio (AR) was not significantly different between longitudinal and transverse analyses in the NF control group, whilst a significant reduction in AR was observed in the transverse analysis of DCM hearts (Fig. 1H). Roundness significantly increased in the transverse analysis of both NF control and DCM (Figure 1I), whilst circularity significantly increased in the transverse nuclei of NF controls but not DCM hearts (Fig. 1J). Solidity was similar between longitudinal and transverse nuclei in NF controls but underwent a significant decrease in the transverse nuclei of DCM hearts (Fig. 1K).

**Figure 1.**
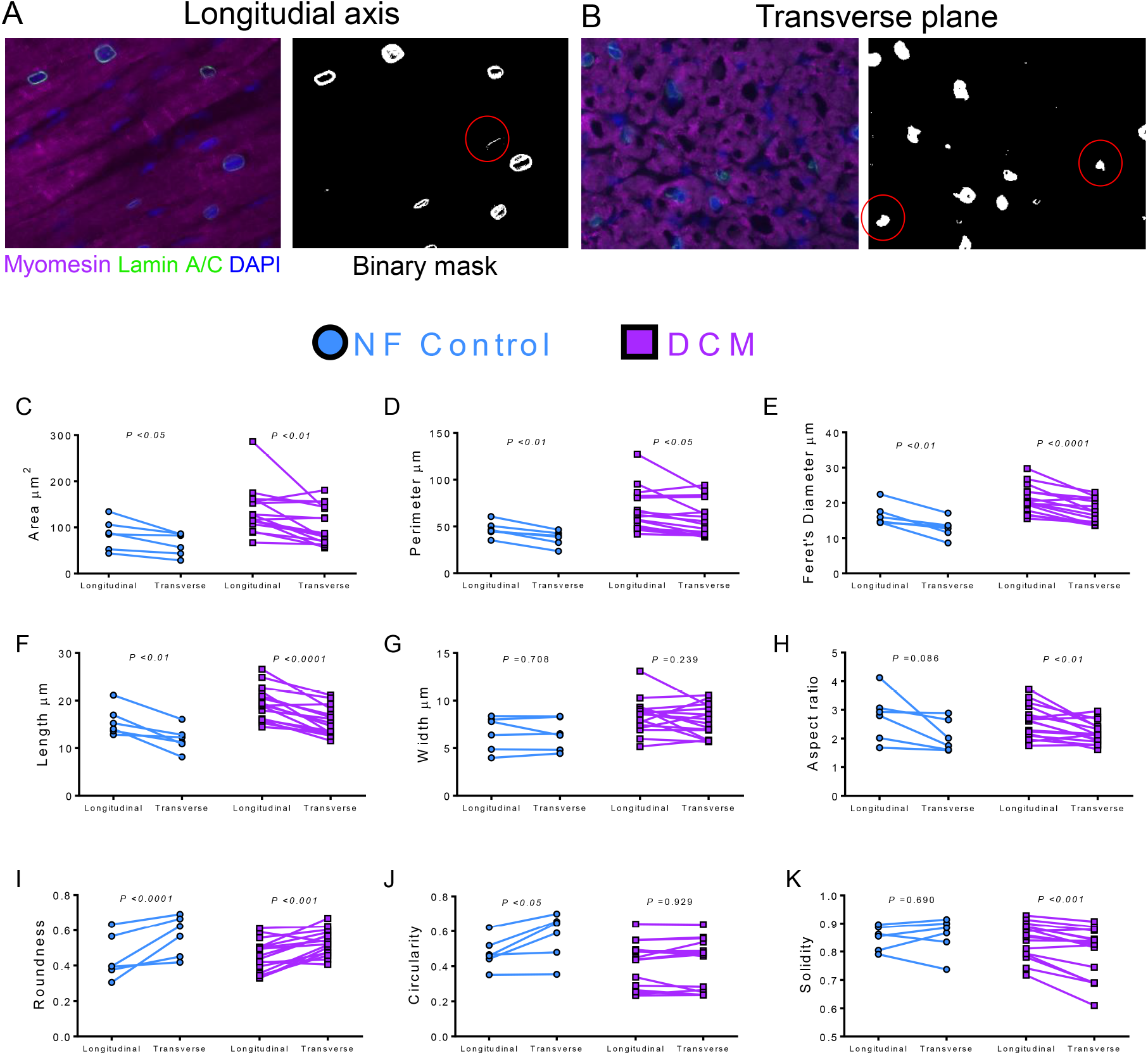
Within-group size and shape analysis of nuclei categorised as longitudinal versus nuclei categorised as transverse revealed discordance in differences exhibited by Dilated Cardiomyopathy (DCM) compared to non-failing (NF) control cardiomyocytes. Representative epifluorescence micrographs and corresponding images subjected to thresholding and segmentation of nuclei in **A**. a section displaying longitudinal nuclei and **B**. a section displaying transverse nuclei. Nuclei circled in red were excluded because of incomplete segmentation (**A**) or did not meet criteria for designation as cardiomyocyte nuclei (**B**). Analysis performed in NF Control and DCM samples for **C**. Area, **D**. Perimeter, **E**. Feret’s diameter, **F**. Length, **G**. Width, **H**. Aspect ratio, **I**. Roundness, **J**. Circularity, and **K**. Solidity. n = 6 NF Control and 16 DCM samples. The range of nuclei counted in each sample was 28 - 92 in the longitudinal axis and 23 - 103 in the transverse axis. Paired two-tailed *t* tests were performed and P<0.05 was considered statistically significant.

The discordance between differences in circularity and solidity of longitudinal and transverse nuclei in NF compared to DCM heart tissue led us to hypothesise that these paramenters may be crucial in determining histopathologic differences relevant for disease, therefore we proceeded to test for differences between NF control and DCM (Table 1). In the longitudinal axis, our analysis confirmed previous reports that nuclei display a larger area in myocardial tissue of DCM patients (6). Concomitant with this, perimeter, length, width and Feret’s diameter were significantly increased in DCM nuclei, whereas DCM nuclei were similar to NF controls with respect to all of the shape characteristics. However, analysis of transverse nuclei showed that most parameters retained nuclear size and shape characteristics with some notable exceptions. Firstly, though average nuclear area was ~40 μm larger in DCM, it was no longer statistically significant, and circularity was significantly decreased in DCM compared to NF controls. When DCMs are compared to NF controls, nuclear solidity was also decreased in transverse DCM (Δ = 0.057) compared to longitudinal DCM (Δ = 0.009), though this did not reach conventionally accepted levels of statistical significance.

We then merged the datasets to perform a combined analysis to yield a dataset that statistically speaking, broadly agreed with the longitudinal dataset conclusion that size parameters were increased, without differences in the shape of DCM nuclei. From these data we conclude that orientation is an important consideration for the morphological analysis of nuclear size and shape in cardiomyocyte nuclei, and that morphological disturbances are more prominent when nuclei are viewed in the transverse plane. The reasons for this could be either technical (transverse nuclei are smaller and morphology defects could be mathematically more sensitive) or mechanistic (mechanical stresses on the lateral versus polar regions are different) in cardiac disease. Because solidity and circularity displayed similar patterns of change between NF Controls and DCM, linear regression of two parameters were performed and confirmed a strong correlation between circularity and solidity in all three analysis subsets (Fig. 2A). This analysis also revealed two distinct morphologically defined subsets: one where nuclear circularity was >0.4, and solidity >0.8; and another where it was <0.4 and solidity <0.8. Whilst the former contained a balanced mix of NF Controls and DCM, the latter contained only one NF Control and six DCM samples (Fig 2A).

**Table 1.** Two-sample testing between nuclei of cardiomyocytes from the left ventricles of non-failing control and dilated cardiomyopathy patients, analysed longitudinally, transversely and with the two datasets combined.

|  | Non-Failing Control |  | Dilated Cardiomyopathy |  | P value* |
| --- | --- | --- | --- | --- | --- |
|  | N = 6 |  | N = 16 |  |  |
| | mean ± S.D. | 95% C.I. | mean ± S.D. | 95% C.I. | |
| **Longitudinal analysis** |  |  |  |  |  |
| Area $\mu\text{m}$ | 85.00 ± 33.51 | 49.83 - 120.20 | 135.50 ± 50.03 | 108.80 - 162.10 | <0.05 |
| Perimeter $\mu\text{m}$ | 47.00 ± 8.33 | 38.26 - 55.74 | 67.22 ± 22.44 | 55.27 ± 79.18 | <0.05 |
| Feret's diameter $\mu\text{m}$ | 16.59 ± 3.11 | 13.32 - 19.86 | 20.78 ± 4.01 | 18.64 - 22.92 | <0.05 |
| Length $\mu\text{m}$ | 15.64 ± 3.05 | 12.44 - 18.84 | 19.33 ± 3.47 | 17.49 - 21.18 | <0.05 |
| Width $\mu\text{m}$ | 6.57 ± 1.81 | 4.67 - 8.47 | 8.38 ± 1.82 | 7.41 - 9.35 | 0.0601 |
| Aspect Ratio | 2.77 ± 0.86 | 1.87 - 3.67 | 2.58 ± 0.577 | 2.28 - 2.89 | 0.6851 |
| Roundness | 0.445 ± 0.126 | 0.313 - 0.577 | 0.453 ± 0.090 | 0.405 - 0.501 | 0.7401 |
| Circularity | 0.476 ± 0.090 | 0.382 - 0.570 | 0.415 ± 0.128 | 0.346 - 0.483 | 0.3966 |
| Solidity | 0.849 ± 0.043 | 0.804 - 0.894 | 0.841 ± 0.063 | 0.807 - 0.874 | 0.9119 |
| **Transverse analysis** |  |  |  |  |  |
| Area $\mu\text{m}$ | 63.82 ± 24.63 | 37.97 - 89.67 | 104.00 ± 41.00 | 82.17 - 125.90 | 0.0709 |
| Perimeter $\mu\text{m}$ | 37.42 ± 8.17 | 28.84 - 45.99 | 58.57 ± 19.07 | 48.4 - 68.73 | <0.05 |
| Feret's diameter $\mu\text{m}$ | 12.86 ± 2.78 | 9.94 - 15.77 | 17.37 ± 3.37 | 15.58 - 19.16 | <0.01 |
| Length $\mu\text{m}$ | 11.91 ± 2.60 | 9.18 - 14.64 | 15.55 ± 3.03 | 13.94 - 17.16 | <0.05 |
| Width $\mu\text{m}$ | 6.46 ± 1.65 | 4.73 - 8.200 | 7.87 ± 1.66 | 6.98 - 8.75 | 0.113 |
| Aspect Ratio | 2.08 ± 0.56 | 1.50 - 2.67 | 2.19 ± 0.38 | 1.99 - 2.39 | 0.3568 |
| Roundness | 0.569 ± 0.113 | 0.450 - 0.688 | 0.530 ± 0.072 | 0.492 - 0.568 | 0.4389 |
| Circularity | 0.571 ± 0.131 | 0.434 - 0.709 | 0.415 ± 0.139 | 0.341 - 0.490 | <0.05 |
| Solidity | 0.856 ± 0.065 | 0.788 - 0.924 | 0.799 ± 0.0874 | 0.753 - 0.846 | 0.113 |
| **Combined analysis** |  |  |  |  |  |
| Area $\mu\text{m}$ | 75.33 ± 29.30 | 44.58 - 106.10 | 119.60 ± 35.42 | 100.70 - 138.50 | <0.05 |
| Perimeter $\mu\text{m}$ | 42.67 ± 7.05 | 35.27 - 50.06 | 62.23 ± 17.94 | 52.67 - 71.79 | <0.05 |
| Feret's diameter $\mu\text{m}$ | 15.00 ± 2.49 | 12.39 - 17.62 | 18.99 ± 3.04 | 17.37 - 20.61 | <0.01 |
| Length $\mu\text{m}$ | 14.05 ± 2.43 | 11.50 - 16.60 | 17.43 ± 2.70 | 15.99 - 18.87 | <0.05 |
| Width $\mu\text{m}$ | 6.46 ± 1.75 | 4.63 - 8.30 | 8.12 ± 1.36 | 7.40 - 8.85 | 0.0601 |
| Aspect Ratio | 2.54 ± 0.67 | 1.83 - 3.24 | 2.40 ± 0.41 | 2.18 - 2.62 | 0.7401 |
| Roundness | 0.491 ± 0.111 | 0.374 - 0.607 | 0.490 ± 0.071 | 0.452 - 0.528 | 0.7964 |
| Circularity | 0.511 ± 0.099 | 0.408 - 0.615 | 0.421 ± 0.127 | 0.353 - 0.488 | 0.0972 |
| Solidity | 0.848 ± 0.053 | 0.792 - 0.903 | 0.826 ± 0.070 | 0.789 - 0.863 | 0.6318 |

\*Determined by a two-tailed Mann-Whitney U test for ranks S.D. = Standard deviation, C.I. = Confidence Interval

A limitation of our study is that our NF control group was relatively small. To ascertain whether a sample was an outlier, future studies should strive to address this problem, notwithstanding the obvious complexity involved in obtaining ‘healthy heart’ control samples. It is also important to note that people cannot be ‘controlled’ like murine experiments. Humans have diverse genotypes and phenotypes, different life stresses, different diets, and exercise to different extents, so when they develop cardiomyopathies, many of which are familial, the analysis will contain parameters that impact on the individual in unpredictable ways. Under these circumstances, the identification of distinct clusters means that linear regression of circularity and solidity can potentially increase the sensitivity of nuclear morphology analysis compared to individual two-sample tests for each parameter, especially in longitudinal and combined analyses. Additionally, visualisation of example nuclei from 3D projections of serially acquired z-planes by confocal microscopy showed that nuclei captured at an angle of 45 degrees between the longitudinal and transverse focal planes are more likely to resemble longitudinal rather than transverse nuclei with respect to shape characteristics (Fig. 2B, suppl. movies 1 & 2). This means that 2D analysis is likely to underestimate the changes to shape characteristics in scenarios where margin calls are made with respect to orientation. Ultimately, this finding suggests that margin calls, in which nucleus orientation is difficult to ascertain, could be categorised as longitudinal to maximise the sensitivity of transverse nuclei analysis, especially when 1) large sample sizes are difficult to attain as in the case of studies involving the study of human heart tissue and 2) in the case that two-sample t tests are the desired statistical method. Furthermore, the potential to perform cardiomyocyte isolations in these types of samples (5) coupled with recent advancements in microscopy (18) may facilitate more precise analysis of nucleus shape in different orientations.

**Figure 2.**
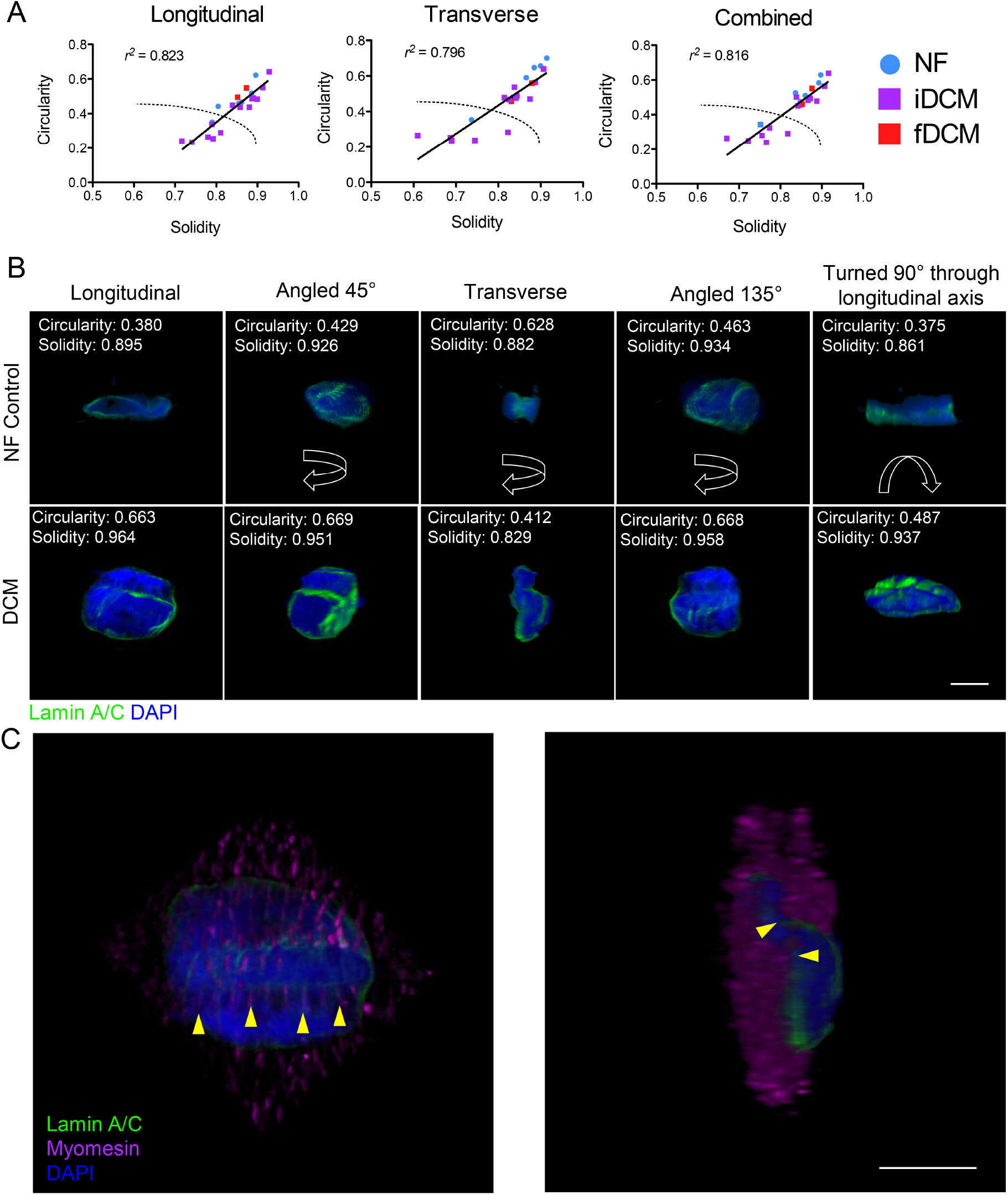
Nucleus shape characteristics circularity and solidity correlate and reveal distinct subclasses of DCM myocardium identifiable in all orientations. **A**. Linear regression of circularity and solidity in longitudinal, transverse and combined analyses showed a strong correlation and two distinct clusters of datapoints visible even in the longitudinal analysis n = 6 NF Control and 16 DCM samples. **B**. Example nuclei from NF Control and DCM tissue sections rotated several angles through Y and then 90° through X to illustrate orientation dependent differences in nuclear shape. **C**. The sarcomeric marker myomesin packed tightly within nuclear in-folds of the same DCM nucleus (yellow arrows). Scales = 10 μm.

### What do nuclear shape defects mean in cardiomyopathy?

Only two of the samples assessed in our study were associated with known mutations responsible for DCM. One patient possessed a mutation in *MYOM1* encoding myomesin and another in *PKP2* encoding plakophilin 2 and both were in the sub-group free of nuclear morphology defects (red squares, Fig 2A). We propose that the latter group may represent a histological sub-classification in which nuclear morphology is disrupted and since determinants of nuclear shape and size are varied (19), this may help to guide and inform on disease processes and mechanism, especially in cases of unexplained/idiopathic cardiomyopathy. For example, abundance of microtubules and the intermediate filament desmin is known to increase in heart failure (20) and its disruption has been shown to induce nuclear morphology shape changes in isolated rat cardiomyocytes, coupled with coordinated changes to the expression of gene sets (16). Moreover, changes in myosins may also drive nuclear shape changes from the outside in with subsequent deleterious effects on nuclear membrane integrity and efficiency of DNA damage repair (21). Accordingly, we were able to observe myofilaments occupying the spaces where nuclei had folded inwards in DCM nuclei (Fig 2C).

Intranuclear mechanisms may also be important, since the nucleoskeleton and chromatin are both mechanically operative and may respond to external stimuli in ways which cause the mis-regulation of discreet regions of DNA with potential effects on transcription, DNA replication and DNA integrity (16, 22-24). Although human heart tissue in the end stage of disease does not represent the most tractable model for studying mechanisms, future studies might look to investigate any correlations that may exist between nuclear shape and ploidy and chromatin state by utilising state of the art labelling and sequencing technologies such as the assay for transposase accessible chromatin sequencing (ATAC-seq) (25). Further, investigating abundance of myofibrillar proteins using mass spectrometry and Western blot and gene expression patterns by way of spatial transcriptomics (26), may help to gain at least some mechanistic insight into an understudied problem. In the long term such work could also provide important diagnostic information into patients with unexplained cardiomyopathy deemed suitable for investigation by biopsy.

## Methods

### Data availability

Source data are available at Figshare https://doi.org/10.6084/m9.figshare.12672950.v1

### Human tissue and study approval

This study was performed on 16 DCM tissue samples from explanted hearts of patients undergoing transplantation and 6 NF Control heart tissue samples from organ donors whose heart was ultimately deemed inappropriate for the intended recipient. This study complies with the declaration of Helsinki. Human DCM specimens were obtained from the Sydney Heart Bank (Hospital Research Ethical Committee approval H03/118; University of Sydney ethical approval 12146) (27) and from Papworth tissue bank in Cambridge, United Kingdom, and used in accordance with ethical guidelines of King’s College London (REC reference 13/LO/1950) and the current United Kingdom law. All patients gave informed consent. Further details of human samples can be viewed in supplementary tables 1 and 2.

### Indirect immunofluorescence

Samples were removed from storage in liquid nitrogen and warmed to -20°C for approximately 1 hour. Tissue was then mounted onto the stage of a cryostat with Optimal Cutting Temperature (OCT) compound (Agar Scientific) and allowed to equilibrate to the ambient temperature of the cryostat for 15 min; for cutting heart tissue, the stage temperature was set to -22°C and the knife was set to -20°C. The sections were then cut to 10 μm thick and mounted onto high quality polylysine or superfrost slides. The sections were allowed to air dry briefly and then stored at -80°C. For staining, sections were allowed to dry completely and fixed by immersion, in a Coplin jar, in pre-chilled 100% methanol at - 20°C for 5 min. Sections were washed 2x5 min in phosphate buffered saline (PBS). Sections were blocked in 3% donkey serum for 1 hour room temperature (RT). Primary antibodies used were lamin A/C, goat polyclonal antibody (N18), Santa Cruz sc-6215, IgG, diluted 1:1000 and myomesin mouse monoclonal (clone B4) IgG (28), diluted 1:100 in blocking solution applied to the sections and incubated in a humidified chamber overnight at 4°C. Sections were washed 3x5 min in PBS and secondary antibody conjugated to a fluorophore was diluted as appropriate, applied to the sections and incubated in the dark in a humidified chamber for 1 hour RT. 4’,6-diamidino-2-phenylindole (DAPI) was added 1:10,000 for 5 min at the end of the incubation for visualisation of DNA. Sections were washed 3x5 min in PBS in the dark and then mounted in mowiol mounting media (Sigma Aldrich) and allowed to dry in the dark overnight.

### Microscopy

Immunofluorescence was analysed using wide-field epifluorescence microscopy (IX81 microscope, Olympus). Images were captured by an experienced myocardial microscopist at 20x lens magnification. Samples were blinded and imaged regions were randomly selected according to the channel corresponding to myomesin staining to avoid unconscious bias and ‘second guessing’ in the channels containing nuclear markers lamin A/C and DAPI. Myomesin is a marker of the sarcomere and was used to help differentiate between fields of view where myocardium predominantly contained myocytes visible in the longitudinal axis as compared to those in a transverse plane of view. The reason this is made possible is because longitudinally oriented cardiomyocytes display a sequence of regularly interspaced striations typical of the sarcomeres in cardiomyocytes (Fig. 1A), whereas the regularity and symmetry of this feature is much less discernible in transverse cardiomyocytes (Fig. 1B). Subjective assessment of the AR of myocytes was also used to help determine the section plane. Confocal images were acquired using a Nikon A1R. Confocal images were processed for 2D and 3D visualisation in Fiji software (29, 30).

### 2D Analysis of Nuclear Morphology

Quantitative analysis was performed using Fiji software (29). Segmentation of nuclei was performed in a semi-automated manner. Separately acquired red, green and blue (RGB) channel images were ‘stacked to RGB’ to create a combined 16-bit flattened image. This image was then subjected to colour thresholding in the RGB colourspace, which facilitated thresholding of each individual channel. Thresholding was performed in the blue (DAPI) and green (lamin A/C) channels to capture nuclei. Segmented regions were then subjected to measurements of size (area, perimeter, length, width and Feret’s diameter) and shape (AR, roundness, circularity, and solidity). Shape descriptors are values derived from equations designed to provide a quantitative assessment of shape and have been designated as ‘feature space analysis’ in previous studies (31). AR (Fig. 3A) represents the relationship between the major axis of the ellipse (length) and the minor axis of the ellipse (width) and is defined by the equation *length + width*. Roundness (Fig. 3B) explains how far from a circle an ellipse is as a product of length and is defined by the equation (4 x *area*) *+* (*n x major axis^2^*). Circularity defines how far from perfect circle a shape is relative to the perimeter (Fig. 3C) and is defined by the equation (*An x area*) *+* (*perimeter*^2^). The key difference between roundness and circularity is that roundness can only tell one how elliptical or oblong a shape is and does not provide data about the smoothness of the circle. Solidity describes the feature known interchangeably in the literature as nuclear morphology defects/blebbing/in-folding/involutions etc. It is derived from the equation *area + area of the convex hull* in which the convex hull is generated by drawing a line between two neighbouring protrusions and measuring the area contained within (Fig. 3D). All assays and analyses were performed at the James Black Centre, King’s College London and in each case the investigator performing the analysis was blinded to group assignment.

**Figure 3.**
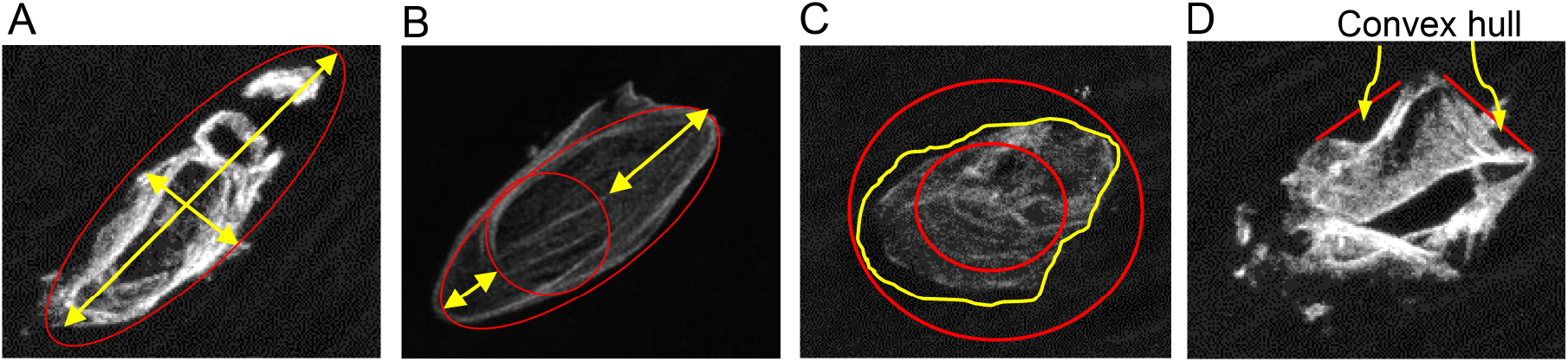
An explanantion of shape descriptors. **A**. A visual explanation of aspect ratio, which describes the length relative to the width **B**. Roundness-which quantifies the how far from a circle an elliptical object is and is relative to length **C**. Circularity, which quantifies the smoothness of a circle, the closer the two red circles are to touching the more circular the shape. If the two circles are superimposed then the value would be ‘1’ i.e. a perfect circle. **D**. Depicts solidity, which quantifies the convex hull (the area created by in-folding) relative to total area of the object.

### Statistics

Analysis of longitudinal versus transverse analyses was performed by paired t-tests. Shapiro-Wilk testing for normality could not be achieved in the NF control group due to the low number of samples therefore non-normal distribution was assumed and differences between NF Control and DCM were tested for by the Mann-Whitney U test for ranks. Data are presented as the mean ± standard deviation (S.D.) with lower and upper 95% confidence intervals (C.I.). *P* values equal to or less than 0.05 were considered significant.

## Data Availability

Source data are available at Figshare https://doi.org/10.6084/m9.figshare.12672950.v1

## Acknowledgements

We would like to acknowledge and thank the Papworth Hospital Research Tissue Bank (Cambridge, United Kingdom) for providing explanted DCM samples. We would also like to thank Dr Andrew Cobb for help with experiment blinding during the course of this study. This study was funded by the British Heart Foundation (grant no. PG/15/93/31834).

## Conflicts of Interest

None to declare

---

Supplemental Tables

**Supplemental Table 1.** Patient information for heart tissue samples of dilated cardiomyopathy patients explanted at transplantation

| Random ID | Biobank centre | Sex | Age range (yr) | Clinical Diagnosis | Gene | Mutation | Ethnicity | Diabetes |
| --- | --- | --- | --- | --- | --- | --- | --- | --- |
| DCM01 | SHB | M | 40-44 | FDCM | NA | NA | Caucasian | No |
| DCM02 | SHB | M | 30-34 | DCM | NA | NA | Caucasian | No |
| DCM03 | SHB | F | 60-65 | FDCM | MYOM1 | E247K | Caucasian | No |
| DCM04 | SHB | M | 50-54 | FDCM IHD | PKP2 | I531S | Caucasian | No |
| DCM05 | Papworth | M | 40-44 | FDCM | NA | NA | Asian british Indian | No |
| DCM06 | Papworth | M | 25-29 | DCM | NA | NA | White british | No |
| DCM07 | Papworth | M | 20-24 | DCM | NA | NA | White british | No |
| DCM08 | Papworth | M | 25-29 | DCM | NA | NA | White British | No |
| DCM09 | Papworth | M | 50-54 | DCM, NYHA class III heart failure | NA | NA | Whit British | No |
| DCM10 | Papworth | M | 60-64 | DCM with biventricular dysfunction | NA | NA | NA | No |
| DCM11 | Papworth | F | 50-54 | Restrictive Cardiomyopathy | NA | NA | White British | No |
| DCM12 | Papworth | M | 35-39 | DCM | NA | NA | NA | NA |
| DCM13 | Papworth | M | 65-69 | DCM with RV ischaemia | NA | NA | NA | No |
| DCM14 | Papworth | M | 30-34 | DCM | NA | NA | White british | No |
| DCM15 | Papworth | M | 40-44 | DCM | NA | NA | White british | Type II |
| DCM16 | Papworth | M | 50-54 | DCM | NA | NA | NA | No |

SHB = Sydney Heart Bank

**Supplemental Table 2.**
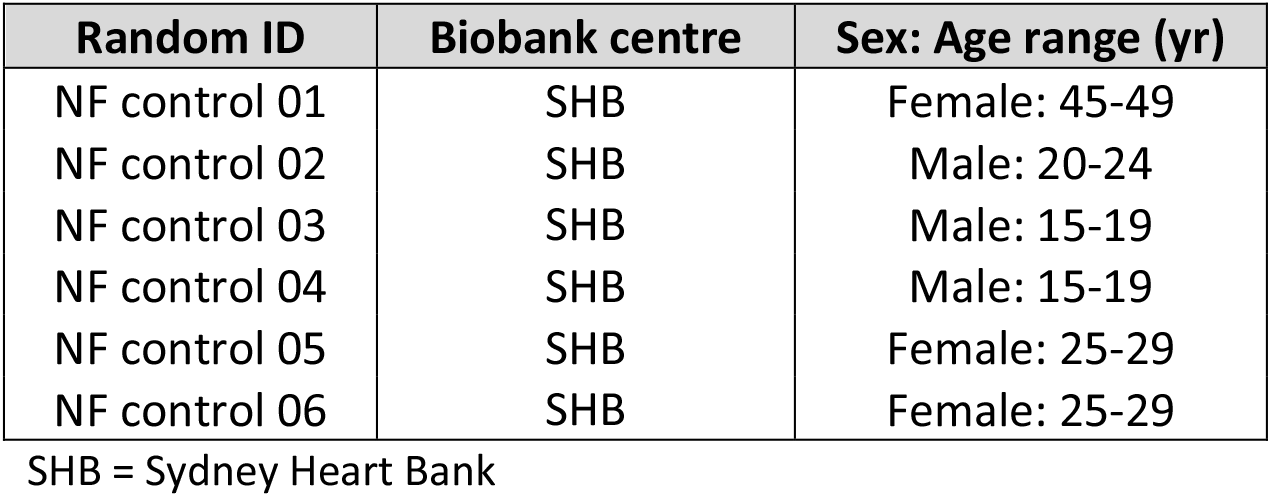
Non-failing controls used in this investigation

Legends for supplemental movies

**Movie 1. 3D stack of a cardiomyocyte nucleus from a non-failing control donor heart**. Myocardial section was stained with lamin A/C antibody (green) and DAPI counterstain (blue). Z-series images were acquired using a 63x objective, deconvolved and reconstructed in ImageJ using the 3D viewer plugin. 360° rotation was recorded and playback was set to 7 frames per second. Movie link: https://doi.org/10.6084/m9.figshare.12672905

**Movie 2. 3D stack of a cardiomyocyte nucleus from a cardiomyopathy patient heart**.Myocardial section was stained with lamin A/C antibody (green) and DAPI counterstain (blue). Z-series images were acquired using a 63x objective, deconvolved and reconstructed in ImageJ using the 3D viewer plugin. 360° rotation was recorded and playback was set to 7 frames per second. Movie link: https://doi.org/10.6084/m9.figshare.12672938

## Notes

### Competing Interest Statement

The authors have declared no competing interest.

### Author Declarations

This study complies with the declaration of Helsinki. Human DCM specimens were obtained from the Sydney Heart Bank (Hospital Research Ethical Committee approval H03/118; University of Sydney ethical approval 12146) and from Papworth tissue bank in Cambridge, United Kingdom, and used in accordance with ethical guidelines of Kings College London (REC reference 13/LO/1950) and the current United Kingdom law. All patients gave informed consent.

